# Molecular epidemiology and antimicrobial resistance phenotype of paediatric bloodstream infections caused by Gram-negative bacteria in Oxfordshire, UK

**DOI:** 10.1101/2021.06.17.21259069

**Authors:** Sam Lipworth, Karina-Doris Vihta, Tim Davies, Sarah Wright, Merline Tabirao, Kevin Chau, Alison Vaughan, James Kavanagh, Leanne Barker, Sophie George, Shelley Segal, Stephane Paulus, Lucinda Barrett, Sarah Oakley, Katie Jeffery, Lisa Butcher, Tim Peto, Derrick Crook, Sarah Walker, Seilesh Kadambari, Nicole Stoesser

**Affiliations:** Nuffield Department of Medicine, University of Oxford, UK; Oxford University Hospitals NHS Foundation Trust, Oxford, UK; Department of Engineering, University of Oxford, UK; NIHR Health Protection Research Unit in Healthcare Associated Infections and Antimicrobial Resistance at University of Oxford in partnership with Public Health England, Oxford, United Kingdom; NIHR Oxford Biomedical Research Centre, Oxford, United Kingdom; Department of Paediatrics, University of Oxford, UK

**Keywords:** neonatal sepsis, paediatric bloodstream infections, whole genome sequencing, antimicrobial resistance

## Abstract

**Objectives:** Gram-negative organisms are common causes of bloodstream infection (BSI) during the neonatal period and early childhood. Whilst several large studies have characterised these isolates in adults, equivalent data (particularly incorporating whole genome sequencing) is lacking in the paediatric population.

**Methods:** We performed an epidemiological and sequencing based analysis of Gram-negative bloodstream infections in children <18 years old between 2008 and 2018 in Oxfordshire, UK.

**Results:** 327 isolates (296 successfully sequenced) from 287 patients were included. The burden of infection was predominantly in neonates (124/327[38%]). Most infections were caused by *Escherichia coli (149/327[46%])*/*Klebsiella* spp. (69/327[21%]) and *Enterobacter hormaechei (34/327[10%])*. There was no evidence of an increasing incidence of *E. coli* BSIs (IRRy 0.96, 95%CI 0.90-1.30, p=0.30) and for *Klebsiella* spp. there was some evidence that the incidence decreased slightly (IRRy 0.91, 95%CI 0.83-1.00, p=0.06). Similarly the proportion of antimicrobial resistant (across all antimicrobial classes evaluated) isolates did not change over time, though we did identify some evidence of sub-breakpoint increases in gentamicin resistance IRRy 1.86, 95%CI 1.33-2.58, p_heterogeneity_=0.002. The population structure of *E. coli* BSI isolates in neonates and children mirrors that in adults with a predominance of STs 131/95/73/69 and the same proportion of O-antigen serotypes covered by the ExPEC-4V vaccine. In most cases there was no evidence of transmission/point-source acquisition and whole genome sequencing was able to refute a previously suspected *Serratia marcescens* outbreak.

**Conclusion:** Our findings support continued use of current empirical treatment guidelines and suggest that O-antigen targeted vaccines may have a role in reducing the incidence of neonatal sepsis.

## Introduction

Gram-negative bloodstream infections (GNBSI) are a common cause of significant morbidity and mortality globally in neonates and young children^1–4^. Their incidence has increased in both the UK and US over the past decade, particularly in very low birth-weight neonates (VLBW, defined as <1500g)^5,6^. Their association with antimicrobial resistance (AMR) has been highlighted by a recent study in the United States of 721 *E. coli* isolates (including 393 bloodstream infection [BSI]-associated isolates), which found high rates of non-susceptibility to commonly used empirical antibiotics including ampicillin (66.8%) and gentamicin (16.8%), as well as an extended beta-lactamase (ESBL) phenotype in 1 in 20 cases^7^. A recent study of 2,483 neonates with culture-confirmed sepsis in low and middle-income countries showed that *Klebsiella* spp. was the predominant pathogen causing multidrug-resistant neonatal sepsis^8^. In Greece, a retrospective observational study in 16 neonatal intensive care units (NICUs) revealed almost half (45%; 36/80) of *Klebsiella* spp. were resistant to either gentamicin or amikacin^9^. The ability of many Gram-negative bacilli (GNB) to readily acquire and exchange genetic material (particularly AMR genes [ARGs]) via mobile genetic elements means that the proliferation of drug-resistant strains remains a constant threat.

The molecular epidemiology of *E. coli* and *Klebsiella* spp. isolates causing invasive infection in adults has been characterised in large sequencing studies.^10,11^ These have demonstrated the emergence of particular AMR-associated sequence types (e.g. *E. coli* ST131)^12^, the genetic homogeneity of isolates causing community and nosocomial onset infections suggesting a common reservoir^13^, the potential for vaccines to play a role in reducing the incidence of these infections^14,15^, and the emerging threat of the convergence of multidrug resistance and hypervirulence in *Klebsiella* spp^16^. To our knowledge, no study to date has systematically evaluated the molecular epidemiology of *E. coli*/*Klebsiella* spp. and other common causes of GNBSI in a paediatric population; published studies focus predominantly on evaluations of outbreaks caused by AMR-associated strains and/or on neonates (see above)^8,17,18^. In this study we therefore aimed to investigate sequencing data from a relatively large collection of sequentially acquired, unselected bloodstream isolates from neonates and children presenting to hospitals in Oxfordshire, UK, over the past decade.

## Methods

### Isolate selection

Oxford University Hospitals NHS Foundation Trust is a large healthcare provider in the South East of England serving a paediatric population of approximately 142,000 across four hospitals (of which two have emergency and acute general paediatric medicine, and one provides all neonatal/paediatric critical care and specialist paediatric services for the region). The microbiology laboratory additionally provides a service to all regional community healthcare providers. All *E. coli* and *Klebsiella* spp. isolates (deduplicated to one morphotype per 90-day period) from Oct-2008 to Nov-2018 collected from blood cultures of patients <18 years old on the day of collection were included in the study. The same selection criteria were applied to other GNB from August-2011 to September-2018 which were excluded from the initial period due to resource limitations. Prior to 2013 antimicrobial susceptibility testing was performed using disk diffusion; after this the Phoenix BD system was used with European Committee on Antimicrobial Susceptibility Testing (EUCAST) breakpoints. Amikacin susceptibility phenotyping was not routinely performed prior to 2013.

### Sequencing procedures

Frozen stocks were sub-cultured onto Columbia blood agar and incubated overnight at 37°C. DNA extractions were performed using the QuickGene DNA extraction kit (Autogen, MA, USA) as per the manufacturer’s instructions (with an additional mechanical lysis step – FastPrep, MP Biomedicals, CA, USA; 6m/s for 40 secs, done twice). Sequencing was performed using Illumina HiSeq 2500/3000/4000/MiSeq instruments as described previously^19^. All sequencing data have been deposited under NCBI accession number PRJNA604975.

### Bioinformatics

*De novo* assembly was performed using Shovill (v1.0.4)^20^. Reads were mapped to sequence type (ST^21,22^) specific references using Snippy (v4.6.0)^23^ (Table S1). For the four *E. coli* STs with the largest number of isolates (i.e. *E. coli* ST131/95/73/69), we created core genome alignments using Snippy-core with the --mask auto setting, padded with the reference base at invariant positions; these whole genome alignments were used as input to Gubbins (v 2.3.4)^24^. Such recombination-corrected phylogenies were also created for *E*.*hormaechei* (the most common non-*E. coli/Klebsiella* species detected in our study) and *S. marcescens* (because there was thought to have been an outbreak in our neonatal intensive care unit in 2016). We also used genomic distances calculated by Mash^25^ (using -k21 -s 100,000 for within-study comparison of isolates and -k21 -s 1000 for comparison of isolates in this study to our collection of sequences of adult bloodstream infection isolates^26^ from the same region over the same time period due to computational feasibility). Annotation against reference databases (VFDB/ResFinder) was performed using ABRicate (v2.3.4)^27^ with genes called as being present if there was ≥80% coverage and DNA identity compared to the reference. Sequence types were predicted using the MLST tool (v2.19)^28^. For *Klebsiella* spp. isolates speciation and virulence gene detection (appendix) was performed using Kleborate (v2.0.4)^29^. Detailed QC metrics and raw Abricate/Kleborate output have been uploaded to Figshare (https://figshare.com/projects/Paediatric_GNBSIs_in_Oxfordshire/135254).

### Definitions

We defined isolates as being likely of neonatal origin if they originated from infants i) in their first 30 days of life, or were ii) on the neonatal intensive care unit, or iii) under the care of a neonatologist on the day the blood culture was taken. For analytical purposes we classified other children as <12 months, 1-4 years, 5-9 years and 10-17 years of age. Early-onset infection was defined as disease within the first 72 hours of life^30^. We calculated a ‘resistance score’ using a previously described method^31^ (the sum of the number of resistance gene categories carried out of amoxicillin, co-trimoxazole, cefotaxime, gentamicin and ciprofloxacin). We further categorised BSIs according to healthcare exposure prior to onset as follows: nosocomial (>48hours after admission to hospital), ‘quasi-nosocomial’ (within 30 days of last discharge), ‘quasi-community’ (31-365 days since last discharge) and community (>365 days since last discharge)^32^. Genetic relatedness in the form of single nucleotide polymorphism (SNP) thresholds definitively associated with transmission is variably defined for the species evaluated; based on recent studies, we considered a threshold of >20 SNVs between isolates as highly unlikely to be representative of a transmission event^33^.

### Epidemiology/statistics

Routinely collected healthcare data were acquired via pseudonymised linkage in the Infections in Oxfordshire Research Database (IORD). IORD has generic Research Ethics Committee, Health Research Authority and Confidentiality Advisory Group approvals (19/SC/0403, 19/CAG/0144) as a de-identified electronic research database. Data on suspected infectious focus (only available for *E. coli/Klebsiella* spp.) were acquired via linked local infection control records which had been submitted to Public Health England as part of the mandatory surveillance programme. Likely source was identified by infectious disease/microbiology physicians using best clinical judgement or designated as unknown where there was significant uncertainty. For each species, we modelled the number of bloodstream infections (BSIs) per year using negative binomial regression, with the total number of paediatric admissions in each year used as an offset to account for changes in the population over time. Only complete years (i.e. excluding 2008 and 2018) were included in this part of the analysis. Median quantile regression was used to model the relationship between age and virulence gene carriage/antimicrobial gene “resistance score”; this analysis was performed in STATA version 15. All other statistical analysis was performed in R v4.0.3^34,35^.

## Results

Microbiology of neonatal and paediatric Gram-negative bloodstream infections in Oxfordshire

Of the 327 GNBSI isolates cultured during the study period from individuals aged <18 years, 149 (46%) were identified as *E. coli* and 69 (21%) as *Klebsiella* spp.; the remaining 109 (33%) belonged to other species (n.b. latter category only collected from 2011). There was no evidence of a change in the incidence of *E. coli* bloodstream infections (BSIs) either overall (incidence rate ratio per year [i.e. the relative increase/decrease in incidence per year] IRRy: 0.96, 95%CI: 0.90-1.03, p=0.30) or in neonates (IRRy: 0.97, 95%CI: 0.86-1.06, p=0.39). This was also the case for other Gram-negative species (i.e. non *E. coli* and *Klebsiella* spp.) both overall (IRRy: 1.07, 95%CI: 0.93-1.24, p=0.33) and in the neonatal group (IRRy: 0.83, 95%CI: 0.65-1.07, p=0.19). Conversely there was some evidence of a decrease in the overall number of *Klebsiella* spp. BSIs in the same period (overall IRRy: 0.91, 95%CI 0.83-1.00, p=0.06; neonates IRRy: 0.80, 95%CI: 0.69-0.92, p=0.002).

Neonates had the highest number of GNBSI (124/327 [38%], of which 8 were early-onset) followed by other infants <1y (66/327 [20%]) (Table 1). There were 62/327 (19%) cases in the 1^st^-4^th^ years of life, 28/327 (9%) in the 5^th^-9^th^ and 47/327 (14%) in the 10^th^-17^th^. A higher proportion of isolates came from males than would be expected from the birth:sex ratio (201/327 [61%], multinomial goodness of fit p <0.001). BSIs in neonates, children aged 1-4 and 10-17 were predominantly nosocomial or quasi-nosocomial whereas in those aged 3-12 months and 5-9 years they were predominantly community or quasi-community onset (Table 1).

**Table 1.** microbiological and patient characteristics of patients with Gram-negative bloodstream infections in Oxfordshire stratified by age of onset. P values represent row-wise comparisons using Fisher exact tests for categorical and Kruskal-Wallis tests for continuous variables.

|  |  | Neonatal<br>Early<br>Onset | Neonatal<br>Late<br>Onset | <12<br>months | 1-4 years | 5-9 years | 10-17<br>years | p |
| --- | --- | --- | --- | --- | --- | --- | --- | --- |
| Species (%) | *Acinetobacter*<br>spp. | 1 (12.5) | 3 (2.6) | 6 (9.1) | 9 (14.5) | 4 (14.3) | 4 (8.5) | <0.001 |
|  | *E. coli* | 6 (75.0) | 51 (44.0) | 42 (63.6) | 17 (27.4) | 6 (21.4) | 27 (57.4) |  |
|  | *Enterobacter*<br>spp. | 1 (12.5) | 24 (20.7) | 8 (12.1) | 13 (21.0) | 5 (17.9) | 4 (8.5) |  |
|  | *Citrobacter* spp. |  | 2 (1.7) |  | 2 (3.2) | 3 (10.7) | 1 (2.1) |  |
|  | *Klebsiella* spp. |  | 28 (24.1) | 10 (15.2) | 16 (25.8) | 6 (21.4) | 9 (19.1) |  |
|  | *Serratia* spp. |  | 8 (6.9) |  |  |  | 1 (2.1) |  |
|  | *Aeromonas* spp. |  |  |  | 3 (4.8) | 1 (3.6) |  |  |
|  | *Proteus* spp. |  |  |  | 2 (3.2) |  |  |  |
|  | *Pantoea* spp. |  |  |  |  | 3 (10.7) | 1 (2.1) |  |
| Age (days) | Median (IQR) | 0.0 (0.0 to<br>1.2) | 20.0 (11.8<br>to 49.2) | 91.0<br>(57.5 to<br>177.5) | 689.0<br>(512.2 to<br>969.8) | 1893.0<br>(1770.0 to<br>2346.2) | 5391.0<br>(3992.5 to<br>6168.5) | <0.001 |
| Sex (%) | Female | 4 (50.0) | 38 (32.8) | 28 (42.4) | 28 (45.2) | 9 (32.1) | 19 (40.4) | 0.51 |
|  | Male | 4 (50.0) | 78 (67.2) | 38 (57.6) | 34 (54.8) | 19 (67.9) | 28 (59.6) |  |

|  |  | **Neonatal**<br>**Early**<br>**Onset** | **Neonatal**<br>**Late**<br>**Onset** | **<12**<br>**months** | **1-4 years** | **5-9 years** | **10-17**<br>**years** | **p** |
| --- | --- | --- | --- | --- | --- | --- | --- | --- |
| Specimen (%)<br>Category | Nosocomial | 0 (0.0) | 86 (74.1) | 7 (10.6) | 18 (29.0) | 0 (0.0) | 15 (31.9) | <0.001 |
|  | Healthcare<br>associated 0-30d | 2 (25.0) | 15 (12.9) | 8 (12.1) | 21 (33.9) | 13 (46.4) | 16 (34.0) |  |
|  | Healthcare<br>associated 31-365d | 0 (0.0) | 3 (2.6) | 29 (43.9) | 11 (17.7) | 4 (14.3) | 5 (10.6) |  |
|  | Community | 6 (75.0) | 12 (10.3) | 22 (33.3) | 12 (19.4) | 11 (39.3) | 11 (23.4) |  |
| Admission<br>Length | Median (IQR) | 14.2 (9.9 to<br>27.5) | 51.7 (20.8<br>to 129.7) | 4.1 (2.5<br>to 7.9) | 8.8 (2.5<br>to 30.6) | 3.1 (2.1 to<br>7.8) | 5.5 (2.9 to<br>28.0) | <0.001 |
| Death Within 30<br>Days (%) | No | 6 (75.0) | 86 (76.1) | 44 (67.7) | 52 (83.9) | 21 (75.0) | 39 (83.0) | 0.30 |
|  | Yes | 2 (25.0) | 27 (23.9) | 21 (32.3) | 10 (16.1) | 7 (25.0) | 8 (17.0) |  |
| CRP | Median (IQR) | 8.2 (0.6 to<br>37.9) | 64.1 (16.1<br>to 150.3) | 99.2<br>(45.5 to<br>125.0) | 50.1<br>(10.1 to<br>122.2) | 16.7 (5.0<br>to 56.8) | 59.2 (13.9<br>to 118.4) | 0.08 |
| Amoxicillin | S | 5 (71.4) | 27 (24.1) | 25 (42.4) | 11 (22.0) | 5 (20.8) | 5 (12.5) | 0.002 |
| (missing=35) | R | 2 (28.6) | 85 (75.9) | 34 (57.6) | 39 (78.0) | 19 (79.2) | 35 (87.5) |  |
| Ceftriaxone | S | 6 (85.7) | 96 (85.0) | 57 (93.4) | 43 (81.1) | 23 (85.2) | 36 (85.7) | 0.46 |
| (missing=24) | R | 1 (14.3) | 17 (15.0) | 4 (6.6) | 10 (18.9) | 4 (14.8) | 6 (14.3) |  |
| Meropenem | S | 8 (100.0) | 115 (100.0) | 62 (100.0) | 61 (100.0) | 28 (100.0) | 43 (97.7) | 0.26 |
| (missing=9) | R | 0 (0.0) | 0 (0.0) | 0 (0.0) | 0 (0.0) | 0 (0.0) | 1 (2.3) |  |
| Ciprofloxacin | S | 7 (87.5) | 110 (95.7) | 62 (100.0) | 55 (91.7) | 27 (96.4) | 39 (88.6) | 0.05 |
| (missing=10) | R | 1 (12.5) | 5 (4.3) | 0 (0.0) | 5 (8.3) | 1 (3.6) | 5 (11.4) |  |
| Amikacin | S | 3 (100.0) | 47 (90.4) | 15 (100.0) | 21 (87.5) | 17 (100.0) | 21 (91.3) | 0.63 |
| (missing=193) | R | 0 (0.0) | 5 (9.6) | 0 (0.0) | 3 (12.5) | 0 (0.0) | 2 (8.7) |  |
| Gentamicin | S | 7 (87.5) | 105 (92.1) | 60 (96.8) | 53 (86.9) | 25 (92.6) | 38 (86.4) | 0.27 |
| (missing=11) | R | 1 (12.5) | 9 (7.9) | 2 (3.2) | 8 (13.1) | 2 (7.4) | 6 (13.6) |  |
| Piperacillin-Tazobactam | S | 8 (100.0) | 105 (92.1) | 56 (93.3) | 48 (85.7) | 23 (88.5) | 37 (86.0) | 0.57 |
| (missing=20) | R |  | 9 (7.9) | 4 (6.7) | 8 (14.3) | 3 (11.5) | 6 (14.0) |  |
| Fosfomycin | S | 1 (100.0) | 45 (93.8) | 10 (90.9) | 14 (87.5) | 11 (84.6) | 18 (94.7) | 0.72 |
| (missing=219) | R |  | 3 (6.2) | 1 (9.1) | 2 (12.5) | 2 (15.4) | 1 (5.3) |  |

In our network of hospitals, amoxicillin and gentamicin are used as empirical treatment for neonatal sepsis. In this patient group, 87/119 (73%) isolates were resistant to amoxicillin and 10/122 (8%) to gentamicin (missing data for five and two isolates respectively); 7/118 (6%) isolates (4/56 *E. coli* and 3/27 *Klebsiella* spp.) were resistant to both. In older children (>1 month), ceftriaxone is the empirical agent used in our setting and in this population 24/183 (13%) isolates were resistant (missing data for 20 isolates). The proportion of resistant isolates appeared broadly stable over time (figure 1/S1). For gentamicin however, there was some evidence that proportion of susceptible isolates with an MIC >1mg/L (IRRy 1.86, 95%CI 1.33-2.58) increased compared to those with an MIC <=1mg/L (IRRy 1.13, 95%CI 1.04-1.22) p_heterogeneity_=0.002 (EUCAST gentamicin breakpoint for resistance >2mg/L). There was no evidence of substantial differences in phenotypic profiles between treating specialties (Figure S2). Likewise, the proportion of resistant isolates was similar for nosocomial vs. community-onset cases for all agents except amoxicillin where nosocomial isolates were proportionally more resistant (99/118 (84%) vs 115/174 (66%), table S2), reflecting the higher burden of *Klebsiella/Enterobacter* spp. BSIs in this patient group (77/126 (61%) vs 101/201 (50%); both these genera are normally considered intrinsically resistant to amoxicllin.

**Figure 1.**
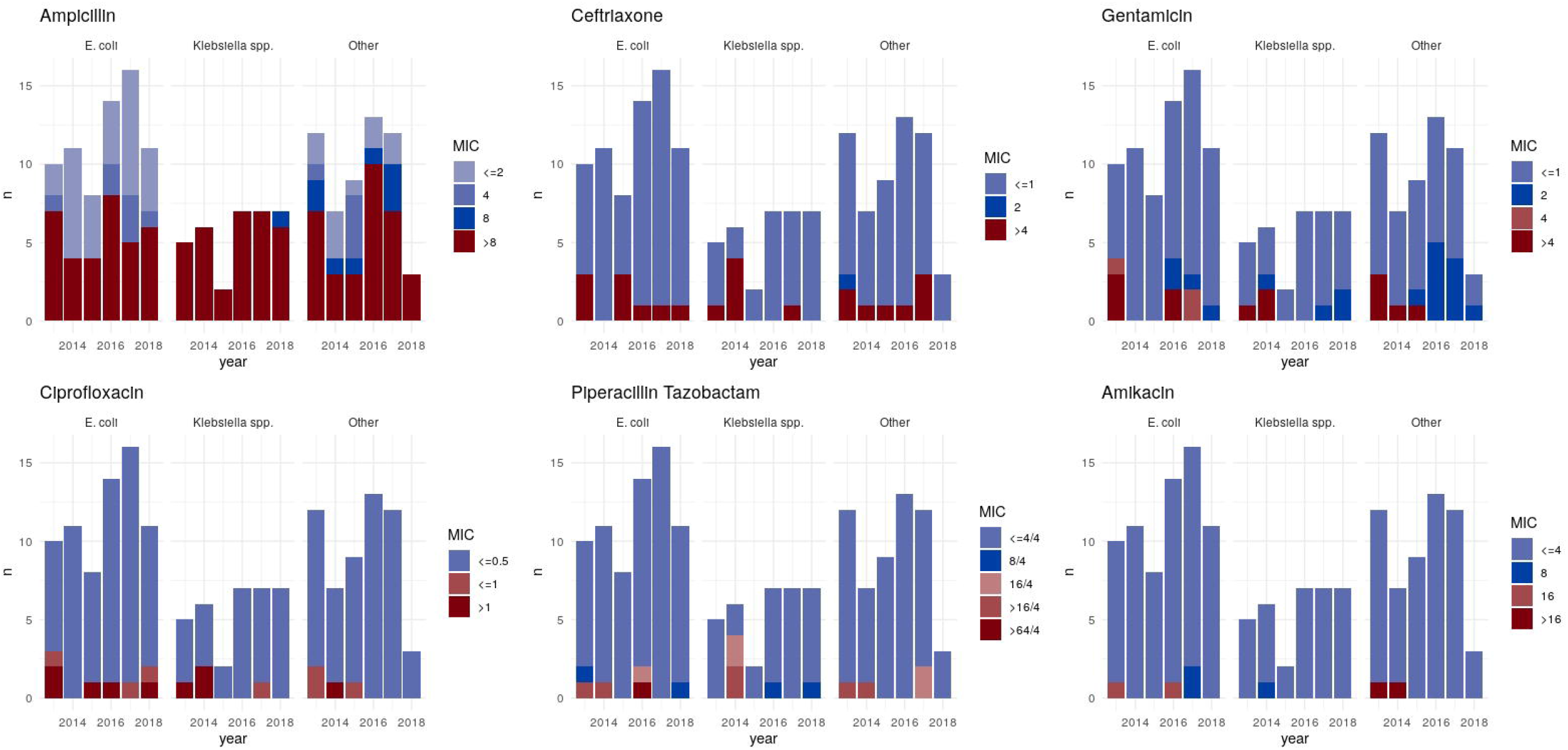
Change in minimum inhibitory concentrations (MIC) of isolates over time.

Data on likely primary infective focus was only available for 78 isolates (66 E. *coli*/12 *Klebsiella* spp.), of these 18 (23%) were of gastrointestinal origin, 8 (10%) from invasive lines, 20 (26%) from the urinary tract, 2 (3%) from the chest and 30 (38%) from unclear/other sources. Seventeen children (six neonates) had infections with >1 species and five (two neonates) had polyclonal infections (>1 ST of the same species).

### Molecular epidemiology of *Escherichia coli* causing BSI, including AMR and virulence gene burden

Of the 149 *E. coli* isolates cultured, 133 (87%) were successfully retrieved for sequencing (Figure 2). The four dominant *E. coli* STs (ST131 n=16, ST95 n=16, ST73 n=20 and ST 69 n=15) were the same as those in adults^26^. There was no evidence that the population structure observed differed from that in the adult population in Oxfordshire (which is also the same as that observed globally^26^; multinomial goodness of fit: p=0.44). The most prevalent O-antigen types were O6 (n=21), O1 (n=14), O2 (n=12), O16 (n=10) and O25 (n=8). The proportion of isolates with an O-antigen included in the ExPEC-4V vaccine^36,37^ (which has been evaluated in a phase-II study in adults), was similar when comparing all children (55/133, 41%) and neonates (25/53, 47%) to that seen in the overall population in our previous study^14^ (1499/3278 (46%); chi-squared: p=0.4 for both).

**Figure 2.**
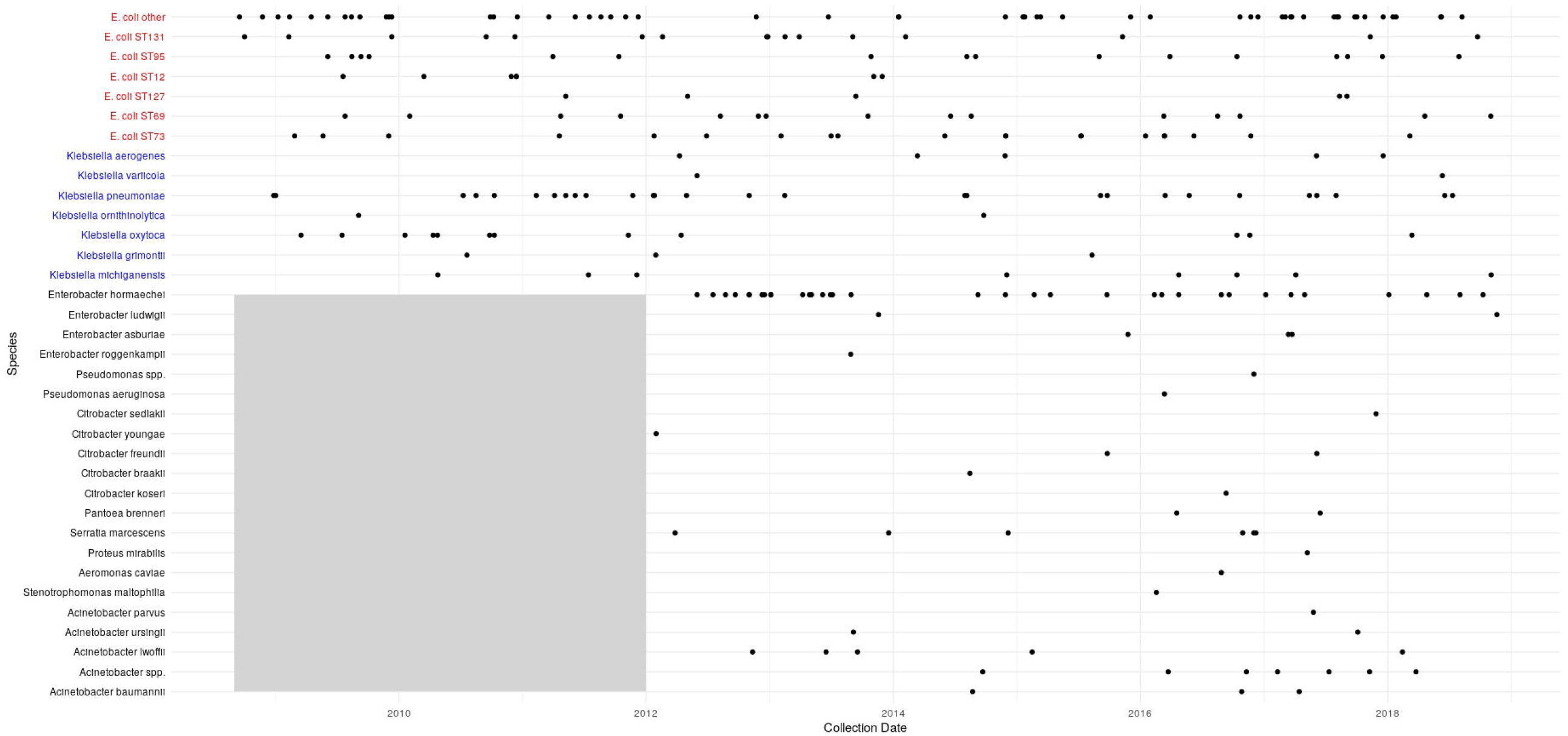
Species (and sequence type, ST, where shown) for sequenced isolate over time. Each point represents the isolation date of a sequenced isolate. The grey shaded area represents the fact that sequencing of non-*E. coli/Klebsiella* spp. isolates did not begin prior to 2012.

Carriage of genes conferring resistance to ampicillin was common (65/133, 49% isolates carrying 82 alleles), with *bla*_*TEM-1B*_ (37/82, 45%) being the most prevalent allele. Eight isolates carried a gentamicin resistance gene (5 *aac(3)*-*IIa* [3 in ST131, 1 each in STs 73 and 23] and 3 *aac(3)*-*IId* [1 each in STs 95, 131 and 69]) and 18/133 (14%) carried a gene conferring resistance to ceftriaxone (mostly ESBL genes, namely *bla*_*CTX-M-15*_ [n=9], *bla*_*CTX-M-14b*_ [n=3] and *bla*_*SHV-102*_ [n=5]); Figure 3). Whilst ST131 was the dominant ESBL-gene-carrying ST (7/16, 44%), other STs carrying these genes included ST73 (4/20), ST2141 (3/3), ST12 (2/7) and STs 23 and 38 (1 each). Quantile regression revealed some association between increasing age and increased resistance score (change in median per year, CMy=0.12, 95% CI 0.04-0.19, p =0.002). Conversely there was some evidence that virulence gene carriage (number of such genes detected by Abricate in the VFDB) was higher in younger children (CMy = -0.53 95%CI -1.03 - -0.033, p=0.04 Figure S3).

**Figure 3.**
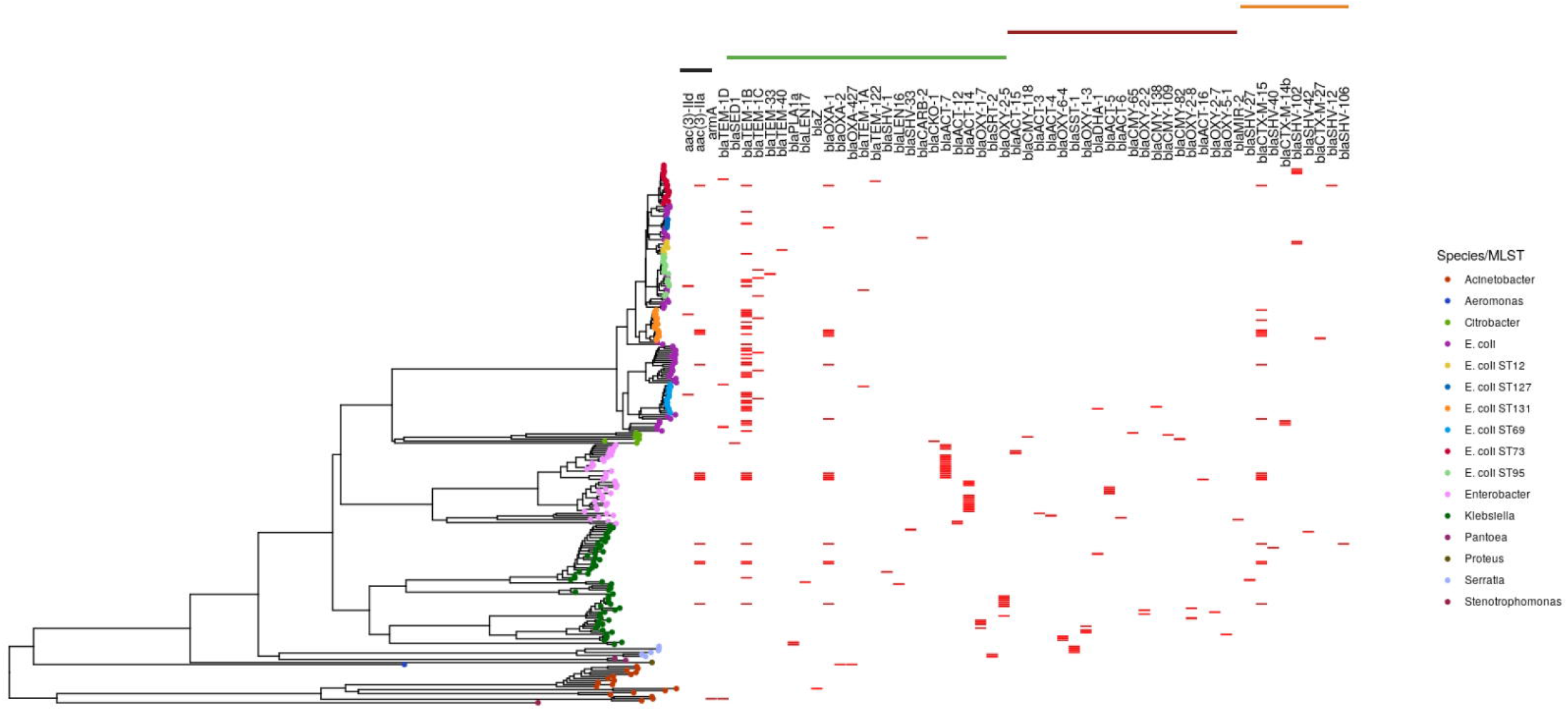
Carriage of antimicrobial resistance genes by isolates sequenced in the study. The tree was created using mashtree and gene presence is shown in red. The black/green/maroon/orange bars at the top represent carriage of genes producing enzymes with activity (as predicted by Resfinder) against aminoglycosides/amoxicillin/cefotaxime (+amoxicillin)/ceftriaxone (+cefotaxime/amoxicillin) respectively.

### Molecular epidemiology of *Klebsiella* spp. causing BSI, including AMR and virulence gene burden

Of the 69 *Klebsiella* spp. isolates cultured, 60 (87%) were successfully retrieved for sequencing. The predominant *Klebsiella* spp. in both neonates and older infants/children was *Klebsiella pneumoniae* (n=28/60 [47%]), though a diverse selection of related species were also occasionally isolated (*K. oxytoca*: n=12 [20%], *K. michiganensis*: n=8 (13%), *K. aerogenes*: 5 (8%), *K. grimontii*: n=3 (5%), *K. ornithinolytica*: n=2 (3%) and *K. variicola:* n=2 (3%)). As we have previously observed in adults and in contrast to the more clonal population seen in *E. coli*, STs causing *Klebsiella* spp. BSI were diverse (n=32 STs in total). There were 11 isolates to which no ST could be assigned. We recently observed that four O-antigens (O2v2, O1v1, O3b and O1v2) were found in 75% isolates from all *K. pneumoniae* BSIs in Oxfordshire over a ten-year period; amongst paediatric BSIs this figure was 61% (17/28; Fisher’s exact test: p=0.15).

Half of *Klebsiella* spp. isolates (30/60) carried the yersiniabactin virulence factor (*ybt*); these were mostly non-*K. pneumoniae* (12/12 K. *oxytoca*, 7/8 K. *michiganensis*, 2/2 K. *ornitholytica* and 3/3 *K. grimontii*). One *K. aerogenes* isolate additionally carried the genotoxin colibactin (*clb*). There was no difference in the proportion of E. coli/Klebsiella spp. isolates carrying a gene conferring ceftriaxone resistance (18/133, 14% vs 7/60, 12%, p=0.82). Only 4/60 (7%) isolates carried a gentamicin resistance gene (four *aac(3)*-*IIa* and one *aac(3)*-*Ia*, similar to *E. coli* (8/133, 6%); Fisher’s exact test: p=0.90).

### Other Gram-negative species

Of the 104 non-*E. coli/Klebsiella* spp. (“other”) GNB cultured in this study, 77 (74%) were successfully sequenced. The predominant species in both neonates and older children was *E. hormaechei* (overall: 34/75, [44%]; neonatal: 14/25 [56%]). This was also the only other species carrying ESBL-producing genes (*bla*_*CTX-M-15*_ in four isolates, Figure 3); these isolates also carried the *bla*_*TEM-1B*_ beta-lactamase as well as *aac(3)*-*IIa* conferring gentamicin resistance. Two isolates carrying class-D beta-lactamases predicted to confer carbapenem resistance were identified (*bla*_*OXA-23*_ in *Acinetobacter baumannii, bla*_*OXA-427*_ in *Aeromonas caviae*).

### Genomic relatedness of isolates

Analysis of recombination-corrected phylogenies for the major *E. coli* STs (131/95/69) demonstrated no clear evidence of direct or temporally related indirect transmission between patients (with the closest genomic distance between patients = 32/87/28 single nucleotide variants [SNV] for ST131/95/69 respectively). There was a cluster of three near-identical ST73 isolates from two patients (3/6 SNPs apart), however the isolation date was ∼2 years apart and these patients had never been admitted to hospital at the same time. There was a cluster of three multidrug-resistant *E. hormaechei* isolates separated by a maximum of 37 SNPs. All three patients had been admitted in the same year but not at the same time and to different wards. Other *E. hormaechei* isolates were genomically diverse (Figure 4). Three *S. marcescens* from the neonatal ICU in 2016 were grouped together in the phylogeny (Figure S4) however the relatively large distances between them (31/81/85 SNPs), made recent transmission unlikely.

**Figure 4.**
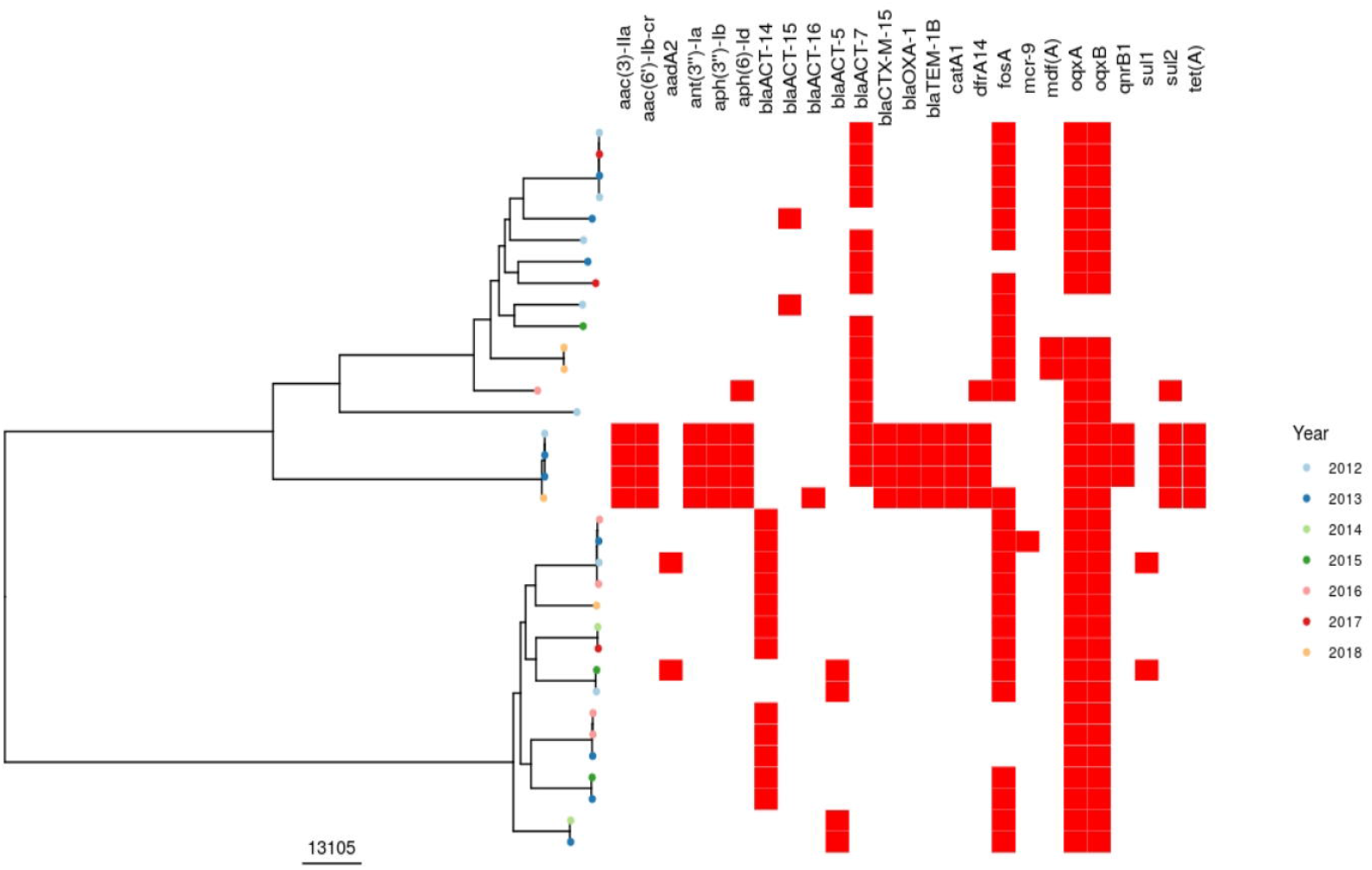
Recombination corrected phylogeny of *Enterobacter hormaechei* isolates sequenced in this study. Tree tip colours show the year of isolates, red bars denote presence/absence of antimicrobial resistance genes. The scale of the tree is shown in SNPs.

There was no difference in the distributions of genomic distances (reflected here by Mash distances) between isolates from patients admitted i) in the same year, ii) to the same ward in the same year, or iii) to the same ward with overlapping admission dates and iv) overall (i.e. all admissions to any ward at any time point) (Kruskal-Wallis rank sum test p=0.30, Figure S5). Finally, when considering *E. coli* and *Klebsiella* isolates in this study, there was no difference between the Mash distances to the nearest paediatric genomic neighbour (median: 0.00093 [IQR: 0.00036-0.0023]) compared to the nearest adult neighbour (median: 0.0011 [IQR: 0.00041-0.0024], Wilcoxon rank sum p=0.33. Figure S6).

## Discussion

*E. coli* was the main causative agent of paediatric and neonatal Gram-negative bloodstream infection in our study over a ten year period. The population structure of *E. coli* isolates causing invasive infection in children mirrors that seen in our centre and globally for adults. We found no evidence of significant nosocomial transmission of isolates causing bloodstream infection, suggesting that in our setting invasive isolates are acquired from the environment, represent colonising gastrointestinal flora^38^, or as a result of transmission from parents/other close contacts. Our findings suggest O-antigen targeted vaccines (currently in phase II/III trials in adults) might have the potential to reduce the incidence of invasive neonatal and paediatric *E. coli* infections, in the former potentially by immunisation of pregnant women. Most isolates remained susceptible to at least one first-line antibiotic justifying continuation of current empirical treatment guidelines which should include an aminoglycoside in neonates^30^. There was however some evidence of increasing (sub-breakpoint) gentamicin MIC which warrants ongoing surveillance, and 1 in 20 GNBSIs were caused by isolates resistant to the ampicillin+gentamicin combination.

The incidence of these infections has remained stable in Oxfordshire over the past decade in children, in contrast to the increase observed in adults. The substantial burden of disease due to paediatric GNBSI occurs in neonates. The absence of evidence of an increasing burden of AMR-associated disease and demonstrable outbreaks likely points to good infection control practice and antimicrobial stewardship, and in our setting we were able to replicate results from a previous study^31^ suggesting greater carriage of virulence genes in *E. coli* isolates from younger children and neonates but higher resistance scores in older children. The latter finding might be explained by increased hospital contact and environmental exposure in this group whereas reasons for and clinical significance of virulence gene acquisition is unclear. Continued epidemiological surveillance with targeted whole genome sequencing to identify strain and/or plasmid outbreaks may help to inform early infection control interventions and would seem particularly justified in these age groups given the high associated morbidity and mortality. In addition we would encourage consideration of clinical trials of existing O-antigen targeted vaccines for prevention of neonatal sepsis.

Our data highlight the importance of *E. hormachei* and as a potential causative agent of neonatal and paediatric sepsis as well as its potential to cause multidrug-resistant disease in this setting. Several small clusters of *E. hormaechei* (part of the *E. cloacae* complex) bloodstream infection in neonates have been described in the literature, with enteral feeding and milk formulations as well as poor general infection control procedures thought to be implicated^39–41^.

Our data highlights the value of whole genome sequencing for the investigation of potential outbreaks on neonatal units. WGS analysis of three isolates from a suspected (on the basis of epidemiology and pulsed-field gel electrophoresis) *S. marcescens* outbreak suggested recent transmission was actually highly unlikely. Had this analysis been performed in real-time it may have reassured clinicians and saved time and resource. Apart from a small cluster of three patients with *E. hormaechei* infections where acquisition from a common source was possible based on genomic and epidemiological analysis (though the patients were never admitted to the same ward), there was relatively high diversity amongst isolates effectively ruling out transmission/point-source acquisition resulting in BSI. However, unwell neonates are often transferred across a geographic region for specialist care and so it is essential that active surveillance mechanisms (collecting and analysing real-time microbiological data) are implemented across networks in order to inform infection prevention measures appropriately.

Notably, whilst we found a lack of clear evidence of transmission between patients in this study, BSIs are likely to represent an extremely insensitive marker of transmission events, as most transmission with these organisms would be expected to lead to colonisation and not necessarily invasive infection. Furthermore whilst the short-read sequencing used in this study allows us to confidently exclude outbreaks of strains, we cannot address the question of potential transmission of AMR genes and virulence factors on plasmids. Additional limitations include the fact that we were not able to sequence all cultured isolates, the relatively sparse availability of data on source attribution and the single region nature of the study. We did not sample other reservoirs potentially contributing to transmission, including the unit environment.

In summary, our study demonstrates a flat overall incidence trend and stable numbers of AMR-carrying GNBSI isolates in children, contrasting with what has been seen in adult populations in the same setting^26^. Pertinently our data supports the ongoing use of an aminoglycoside as part of empirical treatment guidelines in neonatal sepsis. The disproportionate impact of GNBSI on neonates and young infants should encourage active microbiological surveillance across neonatal networks and strategies to prevent disease through vaccine trials in pregnant women.

## Data Availability

Sequencing data has been deposited under project accession number PRJNA604975.

## Ethics Statement

The Infections in Oxfordshire Research Database (IORD; https://oxfordbrc.nihr.ac.uk/research-themes-overview/antimicrobial-resistance-and-modernising-microbiology/infections-in-oxfordshire-research-database-iord/) has generic Research Ethics Committee, Health Research Authority and Confidentiality Advisory Group approvals (19/SC/0403, 19/CAG/0144) which facilitate the pseudo-anonymised linkage of routinely collected NHS electronic healthcare record data from the Oxford University Hospitals NHS Foundation Trust Clinical Systems Data Warehouse and research data (e.g. sequencing data) from the Antimicrobial Resistance and Modernising Microbiology Theme of the Oxford NIHR Biomedical Research Centre, Oxford. IORD links records by a specific, random, number ensuring that no patient-identifiable information is shared with researchers using this resource. We sequenced bacterial isolates from bloodstream infections that are routinely stored by the John Radcliffe Hospital Microbiology laboratory. In the UK, bacterial isolates (such as those sequenced in this study) routinely cultured from human clinical samples do not require ethical approval for analysis under the provisions of the Human Tissue Act as they do not contain any material considered to be human tissue.

