## Supplement for "Molecular epidemiology and antimicrobial resistance phenotype of paediatric bloodstream infections caused by Gram-negative bacteria in Oxfordshire, UK"

**Virulence genes searched for using Kleborate (BLAST hits) in *Klebsiella spp.***

Yersiniabactin (ybt), aerobactin (iuc), salmochelin (iro) and colibactin (clb). Searches were performed using default settings (minimum 90% identity and 80% coverage).

Table S1 – Reference sequences used

| Species (Sequence type) | Reference |
| --- | --- |
| E. coli (131) | HG941718.1 |
| E. coli (95) | NZ_CP012625.1 |
| E. coli (73) | AE014075.1 |
| E. coli (69) | Custom reference, available at 10.6084/m9.figshare.14785854 |
| E. hormaechei | NZ_CP032841.1 |
| S. marcescens | NZ_KQ089767.1 |

Table S2: comparison of phenotypes for community-acquired vs. healthcare-onset bloodstream infection isolates. S – Susceptible, R – Resistant, p values represent Fisher’s exact tests.

|  | **Phenotype** | **Community acquired** | **Healthcare associated** | **p** |
| --- | --- | --- | --- | --- |
| Amoxicillin | S | 59 (33.9) | 19 (16.1) | 0.001 |
|  | R | 115 (66.1) | 99 (83.9) |  |
| Ceftriaxone | S | 162 (87.6) | 99 (83.9) | 0.396 |
|  | R | 23 (12.4) | 19 (16.1) |  |
| Ciprofloxacin | S | 182 (94.8) | 118 (94.4) | 1.000 |
|  | R | 10 (5.2) | 7 (5.6) |  |
| Amikacin | S | 72 (93.5) | 52 (91.2) | 0.743 |
|  | R | 5 (6.5) | 5 (8.8) |  |
| Gentamicin | S | 178 (92.7) | 110 (88.7) | 0.231 |
|  | R | 14 (7.3) | 14 (11.3) |  |
| Piperacillin-Tazobactam | R | 14 (7.5) | 16 (13.2) | 0.117 |
|  | S | 172 (92.5) | 105 (86.8) |  |
| Fosfomycin | R | 5 (8.5) | 4 (8.2) | 1.000 |
|  | S | 54 (91.5) | 45 (91.8) |  |

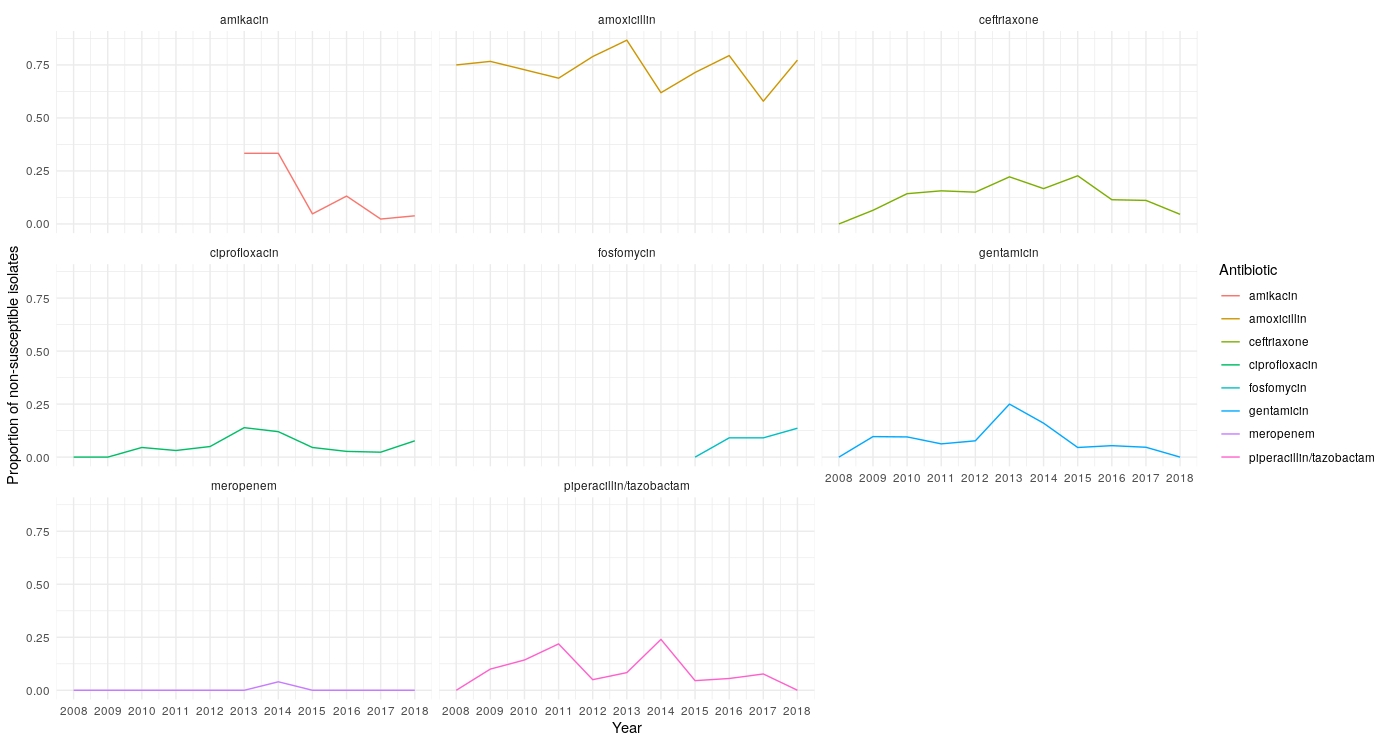

**Figure S1**: Proportion of non-susceptible isolates over time. Routine testing for amikacin and Fosfomycin (2013 and 2015 respectively) started later, hence the blank areas in these plots.

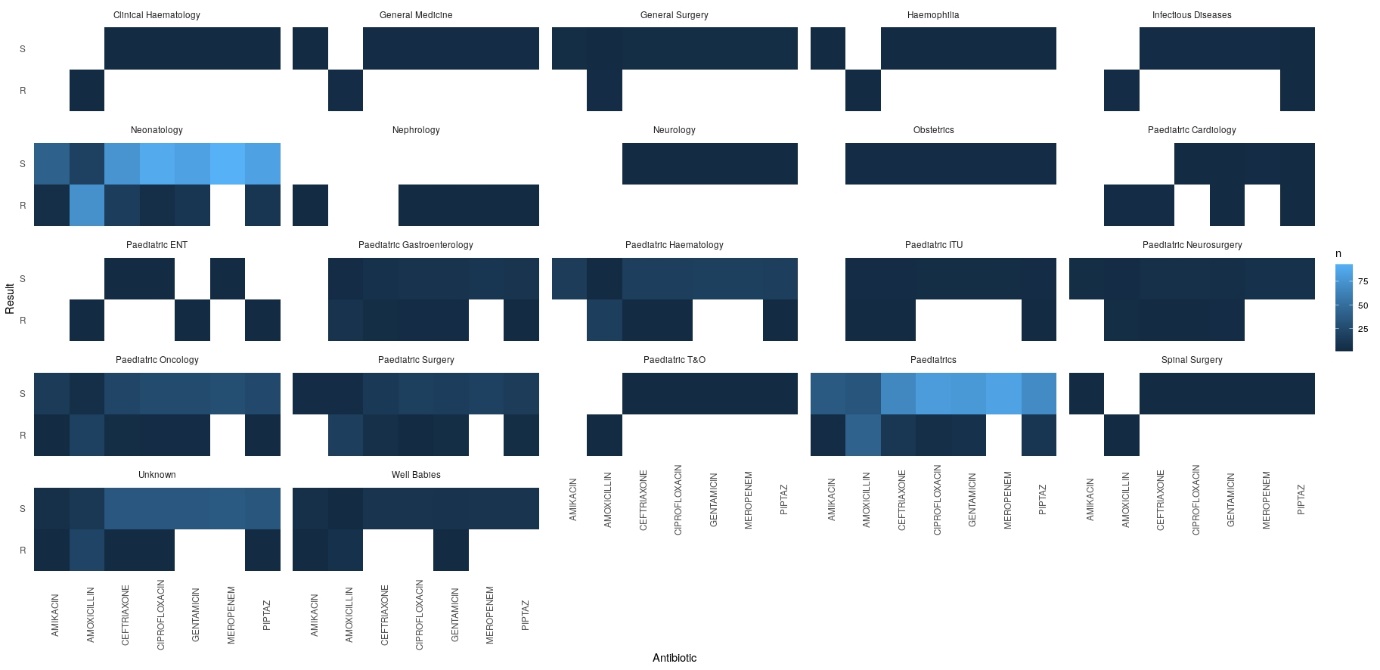

**Figure S2**: Phenotypic profile by primary treatment specialty. Colours represent the number of isolates resistant or sensitive to each antibiotic indicated.

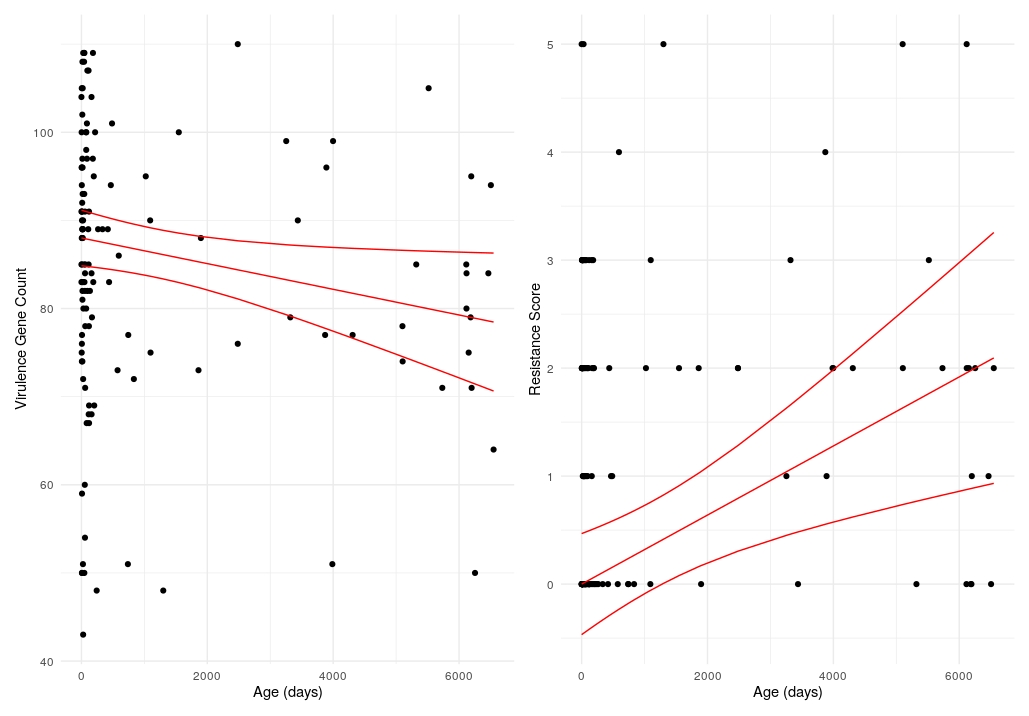

**Figure S3** – virulence gene count (left) and resistance score (right) per isolate against age of host in days. The trend line and 95% confidence intervals were fitted by (median) quantile regression.

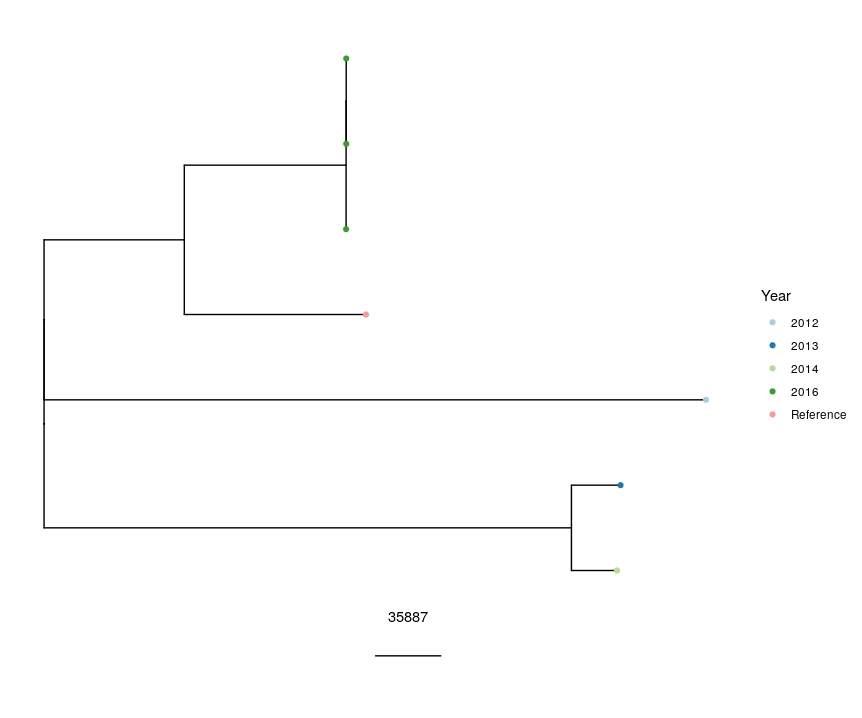

**Figure S4** – Recombination corrected phylogenetic tree of the 6 sequenced *Serratia marcescens* isolates. The scale bar shows distances in SNPs.

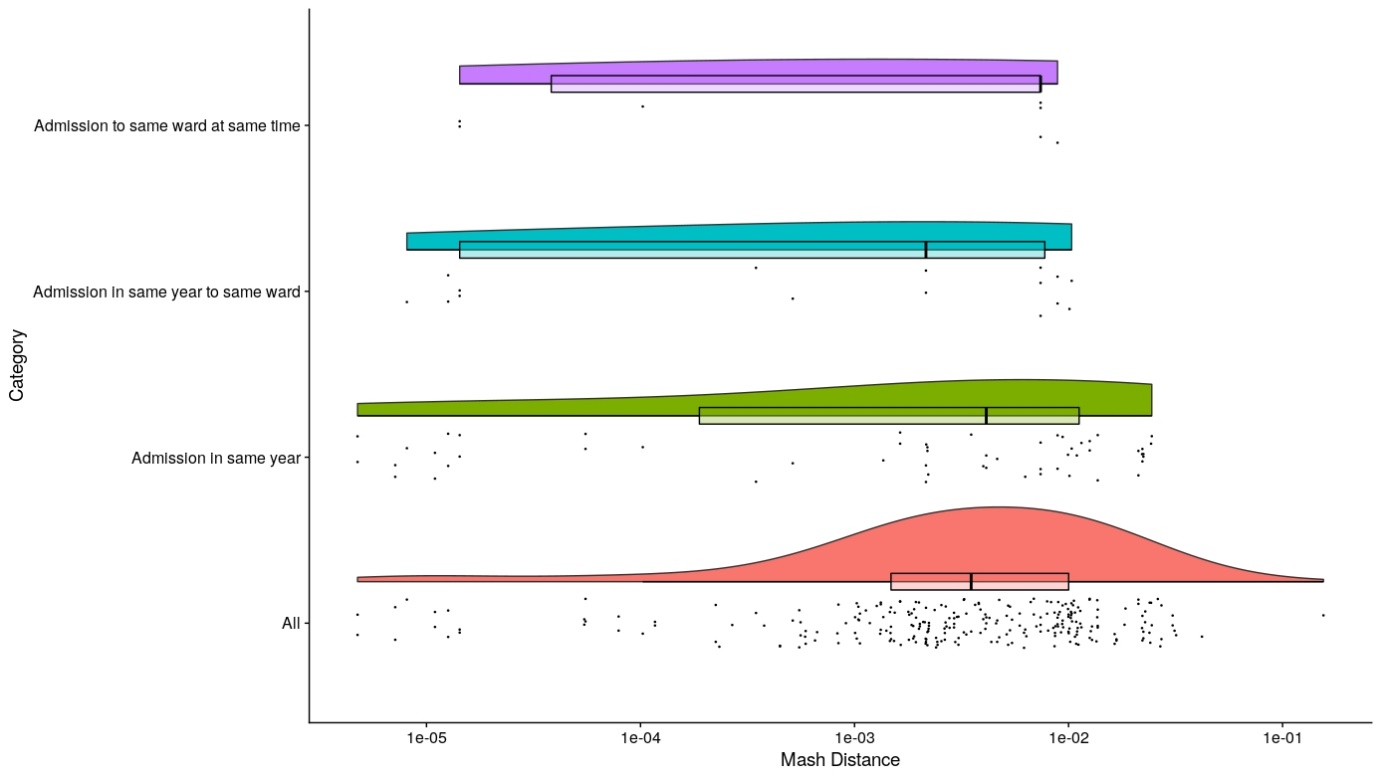

**Figure S5** – rainbow plot showing distributions of Mash distances between isolates from patients with different degrees of epidemiological connections.

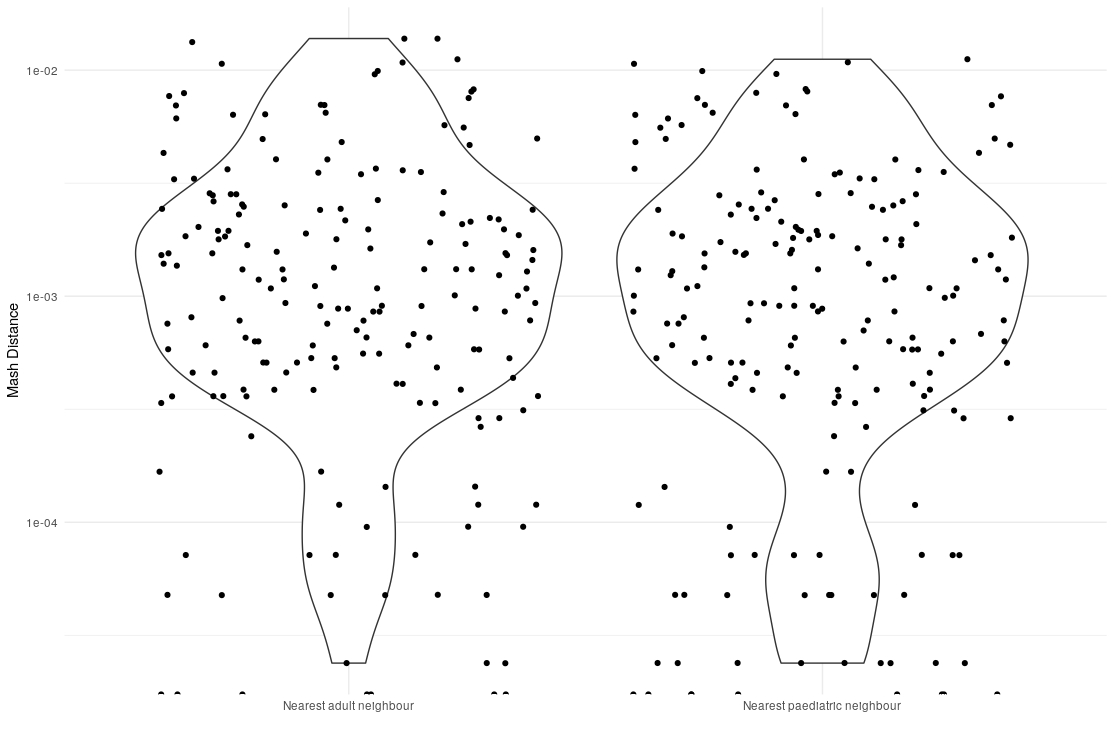

**Figure S6**: Distributions of Mash Distances for the distance to nearest adult neighbour (left) and nearest paediatric neighbour (right). One comparison per patient per group was selected for this analysis.
